# Healthcare Expenditure on Atrial Fibrillation in the United States: The Medical Expenditure Panel Survey 2016-2021

**DOI:** 10.1101/2024.10.30.24316453

**Authors:** Claudia See, Scott Grubman, Nisarg Shah, Jiun-Ruey Hu, Michael Nanna, James V. Freeman, Karthik Murugiah

**Affiliations:** Division of General Internal Medicine, University of California, San Francisco, CA; Division of General Internal Medicine, The Mount Sinai Hospital, New York, NY; Yale School of Medicine, New Haven, CT; Department of Cardiology, Smidt Heart Institute, Cedars-Sinai Medical Center, Los Angeles, CA, USA; Section of Cardiovascular Medicine, Department of Internal Medicine, Yale School of Medicine, New Haven, CT; Center for Outcomes Research and Evaluation, Yale New Haven Health Services Corporation, New Haven, Connecticut, USA

**Author notes:** **Financial Support:** Dr. See received support from the Yale School of Medicine 2023 Fellowship for Medical Student Research. Dr. Murugiah received support from the National Heart, Lung, and Blood Institute of the National Institutes of Health (under award K08HL157727). Dr. Nanna reports research support from the American College of Cardiology (George F and Ann Harris Bellows Foundation), the Patient-Centered Outcomes Research Institute, the Yale Claude D. Pepper Older Americans Independence Center (P30AG021342), and the National Institute on Aging/National Institutes of Health (GEMSSTAR award: R03AG074067). **Relationship with Industry:** Dr. Freeman reports research support from the NIH/NHLBI and the American College of Cardiology National Cardiovascular Data Registry and advisory board/consulting with Boston Scientific, Medtronic, Abbott, Biosense Webster, and PaceMate. Dr. Nanna reports consulting from HeartFlow, Inc., Novo Nordisk, and Merck. **Address for Correspondence:** Claudia See, MD MBA, 1239 4th Ave, San Francisco, CA 94122.

## Abstract

**Objectives:** To provide a contemporary nationally representative assessment of atrial fibrillation (AF) and atrial flutter (AFL) expenditures in the United States.

**Background:** AF prevalence is rising over time and management is evolving. However, there has been no contemporary national assessment of expenditures of AF.

**Methods:** Using Medical Expenditure Panel Survey (MEPS) 2016-2021 data, we identified individuals with AF or AFL using International Classification of Disease (ICD)-10 codes and reported total and categorized expenditures. Using two-part and gamma regression models, respectively, we estimated the incremental expenditures with AF for the entire population and for individuals with common coexisting comorbidities. Among individuals with AF, we identified characteristics associated with higher expenditures.

**Results:** Of a weighted surveyed population of 248,067,064 adults, 3,564,763 (1.4%) had AF. Mean age was 71.9 ± 10.6 years and 45.7% were female. Mean unadjusted annual total healthcare expenditure for individuals with AF was $25,451 ± $1,100 compared with $9,254 ± $82 for individuals without AF. The highest spending categories were inpatient visits ($7,975 ± $733) and prescriptions ($6,505 ± $372). AF expenditures increased over the study period by 11.1%. After adjustment, the incremental annual expenditure attributable to AF was $6,188 per person. Incremental expenditures with AF were highest for those with cancer ($11,967, 95% CI $4,410 - $19,525), while AF did not significantly increase expenditures in HF (-$2,756, 95% CI -$10,048 - $4,535). Modified Charlson Comorbidity Index of 1 or ≥2, uninsured status, cancer, poor income level, ASCVD, COPD, and later survey year were associated with higher expenditures.

**Conclusion:** AF is associated with substantial healthcare expenditures which are increasing over time. With changes in screening and management, expenditures need periodic reassessments.

**CONDENSED ABSTRACT:** *Introduction:* AF prevalence is rising, but contemporary national expenditure assessments are lacking.

*Methods:* Two-part and gamma regression models of MEPS 2016-2021 data (all-payer cross-sectional US survey) estimated AF effect on healthcare expenditures.

*Results:* Of 248,067,064 adults, 1.4% had AF, with mean age 72.1 years and 45.7% female. Mean unadjusted annual expenditure was $25,451 ± $1,100 ($9,254 ± $82 without AF). Incremental adjusted annual expenditure attributable to AF was $6,188. Modified CCI of 1 or ≥2, uninsured status, cancer, poor income level, ASCVD, COPD, and later survey year were associated with higher expenditure.

*Discussion:* AF expenditures are substantial and increasing.

## INTRODUCTION

Atrial fibrillation (AF) is the most common cardiac arrhythmia, affecting between 2.7 to 6.1 million people in the United States (US) (1). Over time, with an aging population and potentially increased detection due to improved and more widespread monitoring devices, the prevalence of AF has been rising and the prevalence is estimated to be 12.1 million by 2030 (2).

AF is associated with significant morbidity, including hospitalizations and risk for cardiovascular and cerebrovascular events (3,4). Given these characteristics, AF is associated with high healthcare expenditures (5,6). The management of AF has been evolving, with increased use of antiarrhythmics and procedures to maintain sinus rhythm as well as increased use of direct-acting oral anticoagulants (DOACs) and left atrial appendage occlusion for stroke prevention, which can also affect expenditures. However, contemporary studies of US national healthcare expenditures for AF care have been lacking. Further, AF frequently co-exists with other important cardiac and non-cardiac comorbidities such as heart failure (HF), atherosclerotic cardiovascular disease (ASCVD), chronic obstructive pulmonary disease (COPD), and cancer, all of which may complicate their management (7-9). How contemporary expenditures of these conditions are influenced by co-existent AF has not been well evaluated.

Accordingly, the aim of this study was to report current annual healthcare expenditures and recent trends for individuals with AF (categorizing expenditures across medical service categories such as hospitalizations and prescriptions), estimate the incremental expenditures nationally attributable to AF, and identify the characteristics of individuals with AF who experience higher expenditures.

## METHODS

### Data Sources

The Medical Expenditure Panel Survey (MEPS) is an annual cross-sectional survey of US families and individuals by the Agency of Healthcare Research and Quality (AHRQ). It is the largest all-payer nationally representative annual cross-sectional survey of medical expenditures of the US civilian, noninstitutionalized population (10). MEPS collects data on demographic characteristics, health conditions, health status, use of medical services, charges and source of payments, access to care, satisfaction with care, health insurance coverage, income, and employment. Overall response rates in the years 2016 to 2021 were approximately 22-46% (11).

The panel design of the survey includes 5 rounds of interviews covering 2 full calendar years. Every year, a new panel of about 15,000 sample households is selected. The set of households selected for each panel is a subsample of households participating in the previous year’s National Health Interview Survey (NHIS) conducted by the National Center for Health Statistics (NCHS).

Data from the MEPS Household Component Full-Year Consolidated Data Files and Medical Conditions Files were pooled across the years 2016 to 2021 and then merged using two variables—patient identifier variable ‘DUPERSID’ and survey ‘Year.’ The study was limited to 2016 and after because the ICD-9 codes used in MEPS data before 2016 were limited to three-digits for confidentiality reasons and could not distinguish AF from other cardiac arrhythmias. For 2016 and 2017, the two-digit panel number from variable ‘PANEL’ was added in front of all DUPERSIDs per the recommendation of AHRQ (12) to be consistent across all survey years. Families and individuals were assigned weights based on demographic proportions in the overall US population (13). The NHIS sample design underwent several changes between 2016-2021, such as discontinuation of oversampling of Asian, Black, and Hispanic minority groups (14) in favor of oversampling of Veterans and middle-sized states (15,16). To compensate for low response rates during the COVID-19 pandemic, MEPS extended the duration of its survey panels by one year (11).

As all data and materials are de-identified and publicly available online at https://meps.ahrq.gov/mepsweb/, this study was exempt from institutional review board approval by Yale University.

### Study Population

The study population included all adults aged 18 years old or older. AF and atrial flutter (AFL) was identified using the International Classification of Disease (ICD)-10 code “I48.” Similarly, ASCVD, HF, COPD, and cancer were identified using appropriate ICD-10 codes (see Supplemental Table 1). The weighted population size was estimated as the average annual population size for the pooled period, based on the recommendation from AHRQ on pooling (17).

### Variables and Pre-Processing

Age was categorized into 18-44, 45-64, 65-84, and ≥85 years old, and sex as male or female. Race/ethnicity was categorized as non-Hispanic White, Hispanic, non-Hispanic Black, and non-Hispanic Asian and other/mixed race. Marital status was categorized as married, never married, and divorced/widowed/separated. Insurance status was categorized as private, Medicare, other public, or uninsured. Education was categorized as no degree, general educational development (GED) or high school (HS) graduate, bachelor’s degree, master or doctorate degree, and other degree. Family income was categorized as poor (families with income less than or equal to the federal poverty line [FPL]), near-poor (100-125% of the FPL), low income (125-200% of the FPL), middle income (200-400% of the FPL), and high income (>400% of the FPL) per the MEPS poverty status variable. In the multivariable models, age and survey year were treated as continuous variables. The Stata ‘charlson’ package was used to calculate the Charlson Comorbidity Index (CCI) score (18). A higher score represents more comorbid conditions.

Total medical expenditure was defined as the sum of direct payments for care across 8 medical service categories: inpatient hospitalization stays, outpatient and office-based visits, prescription medication, emergency department (ED) visits, dental visits, home healthcare, and others (including dental and vision expenditures). Payments were combined across 10 payers such as private insurance, Medicare, Medicaid, Tricare, out-of-pocket, and other. All expenditures and incomes in our analyses were inflation adjusted to year 2021 using the Consumer Price Index (CPI). Home health expenditures were derived by summed home health agency and home health non-agency expenditure MEPS variables. Other expenditures were derived by summed dental, vision, and ‘other’ expenditure MEPS variables.

### Statistical Methods

Continuous variables were presented as mean ± standard deviation (SD) and categorical variables as proportions. Continuous and categorical variables were compared using the Wilcoxon rank-sum test and chi-squared tests, respectively. A two-sided p-value of <0.05 was used for statistical significance. MEPS survey weights were applied using the ‘svy’ function in Stata. Statistical analysis was conducted using Stata Statistical Software: Release 18 (College Station, TX).

### Modeling Healthcare Expenditure

For expenditure data which are typically right-skewed (19), a two-part model is commonly used—the first model is a logistic or probabilistic (probit) regression model which estimates the probability of zero versus positive expenditures, and the second model, conditional on having a positive annual healthcare expenditure, such as a generalized linear model with gamma distribution and a logarithmic-link function that estimates the average expenditure per capita (20,21).

Accordingly, to estimate incremental total healthcare expenditure attributable to AF, a two-part regression model was created using the entire pooled population with and without AF. Covariates included in multivariable adjustment were AF, age, sex, race/ethnicity, survey year, CCI, poverty level, education status, marital status, and insurance status. National estimates of AF expenditure were extrapolated by multiplying the incremental expenditure per person by the survey weighted number of individuals with AF in the US in our study.

Second, incremental expenditures attributable to AF within subpopulations of individuals with 4 other common chronic comorbidities of cancer, ASCVD, COPD, and HF were estimated by creating a gamma regression model for each subpopulation and calculating average marginal effect with 95% confidence intervals (CI). Gamma models were used instead of two-part models because of the low prevalence of individuals with no expenditures within the comorbidity subpopulations. For these gamma regression models, a ‘modified’ CCI was used by excluding the 4 comorbidities of ASCVD, cancer, HF, and COPD (22), resulting in 3 final score categories of 0, 1, and ≥2. This was done to enable inputting these comorbidities as separate covariates into the model to study their individual effects.

Third, individual characteristics associated with increased total healthcare expenditures were determined by creating a gamma regression model only for individuals with AF and calculating odds ratios (OR) with 95% CI. For this gamma regression model, the ‘modified’ CCI covariate was also used.

## RESULTS

### Study Population Characteristics

The average annual surveyed population was 22,911 US adults aged ≥18 years. Of these, 98.6% had complete data (n=22,583), equivalent to a survey weighted population of 248,067,064 adults. Within this population with complete data, 1,902 survey respondents had AF, equivalent to a survey weighted population of 3,564,763 adults and an overall AF prevalence of 1.4%.

Table 1 shows demographics and comorbidities of the overall population, with and without AF. Among individuals with AF, the mean age was 71.9 ± 10.6 years, 45.7% were female, and 89.0% were non-Hispanic white. The median CCI was 1 (Interquartile Range [IQR] 0-1). Among individuals without AF, the mean age was 47.5 ± 18.2 years, 51.8% were female, and 62.3% were non-Hispanic white. The median CCI was 0 [0-0] for individuals without AF. The prevalence of ASCVD between individuals with and without AF was 34.7% vs. 5.2%, cancer 12.6% vs. 3.4%, COPD 9.5% vs. 2.2%, and HF 7.0% vs. 0.6% respectively (all p<0.001).

**Table 1:**
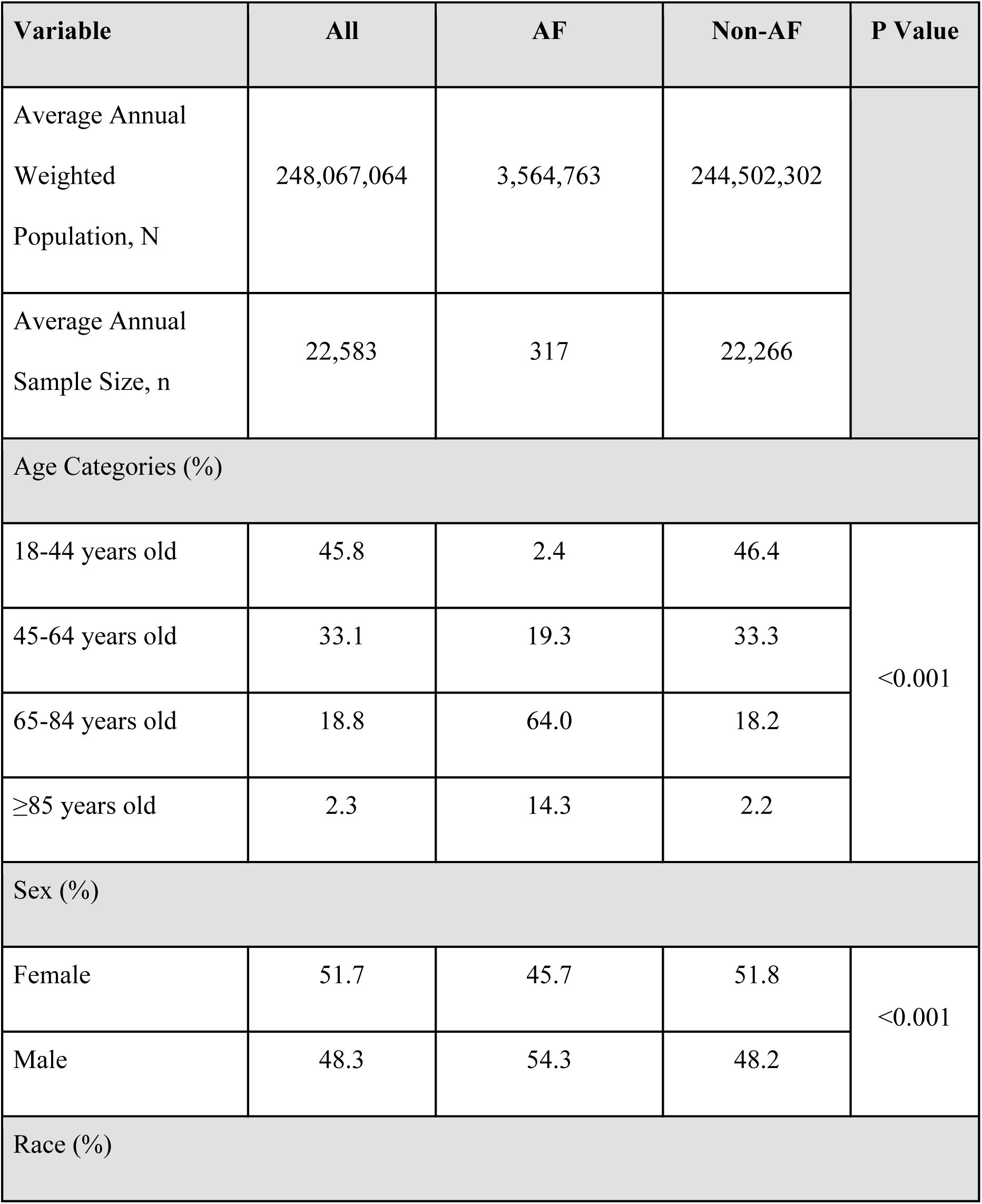
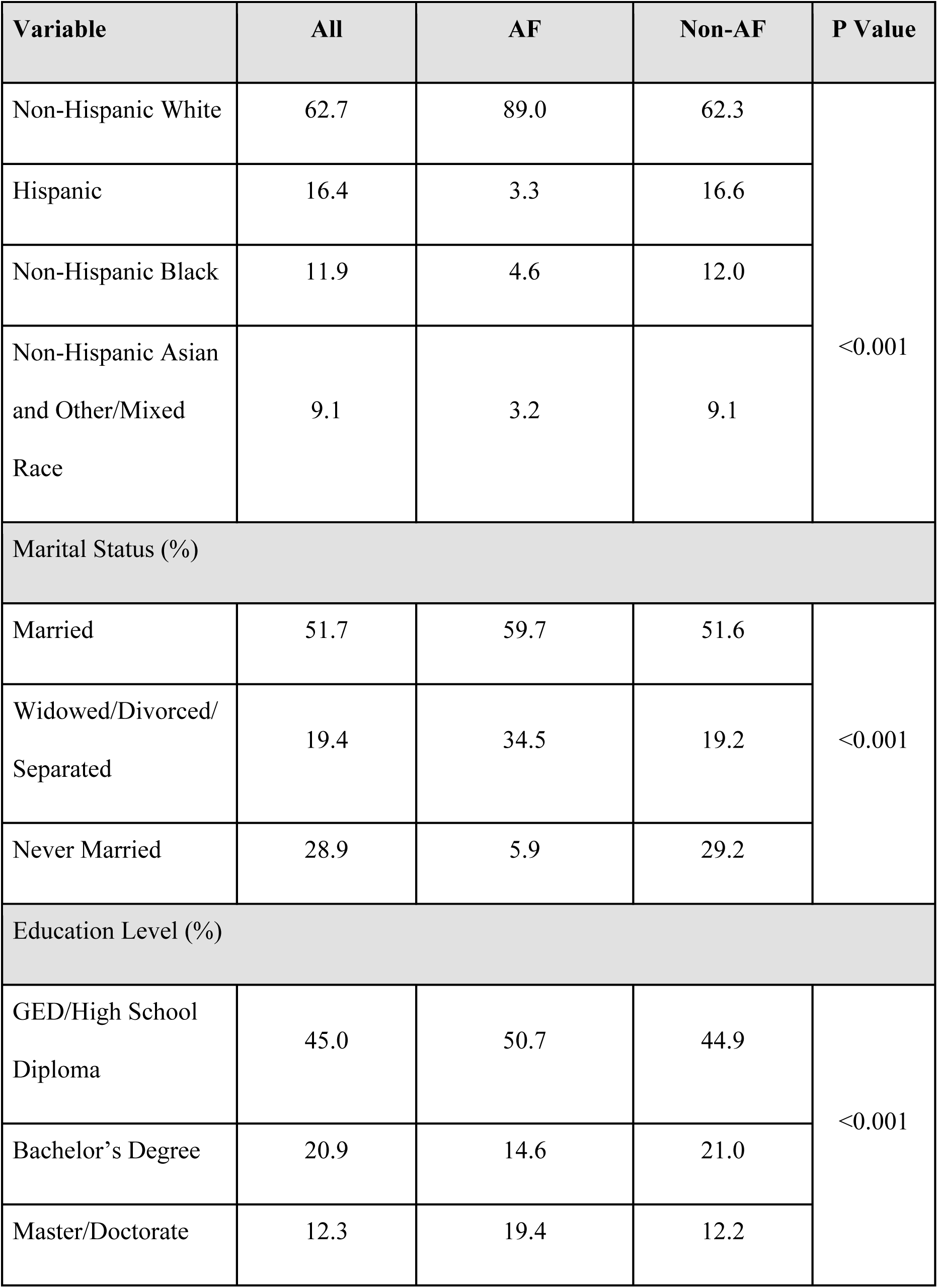
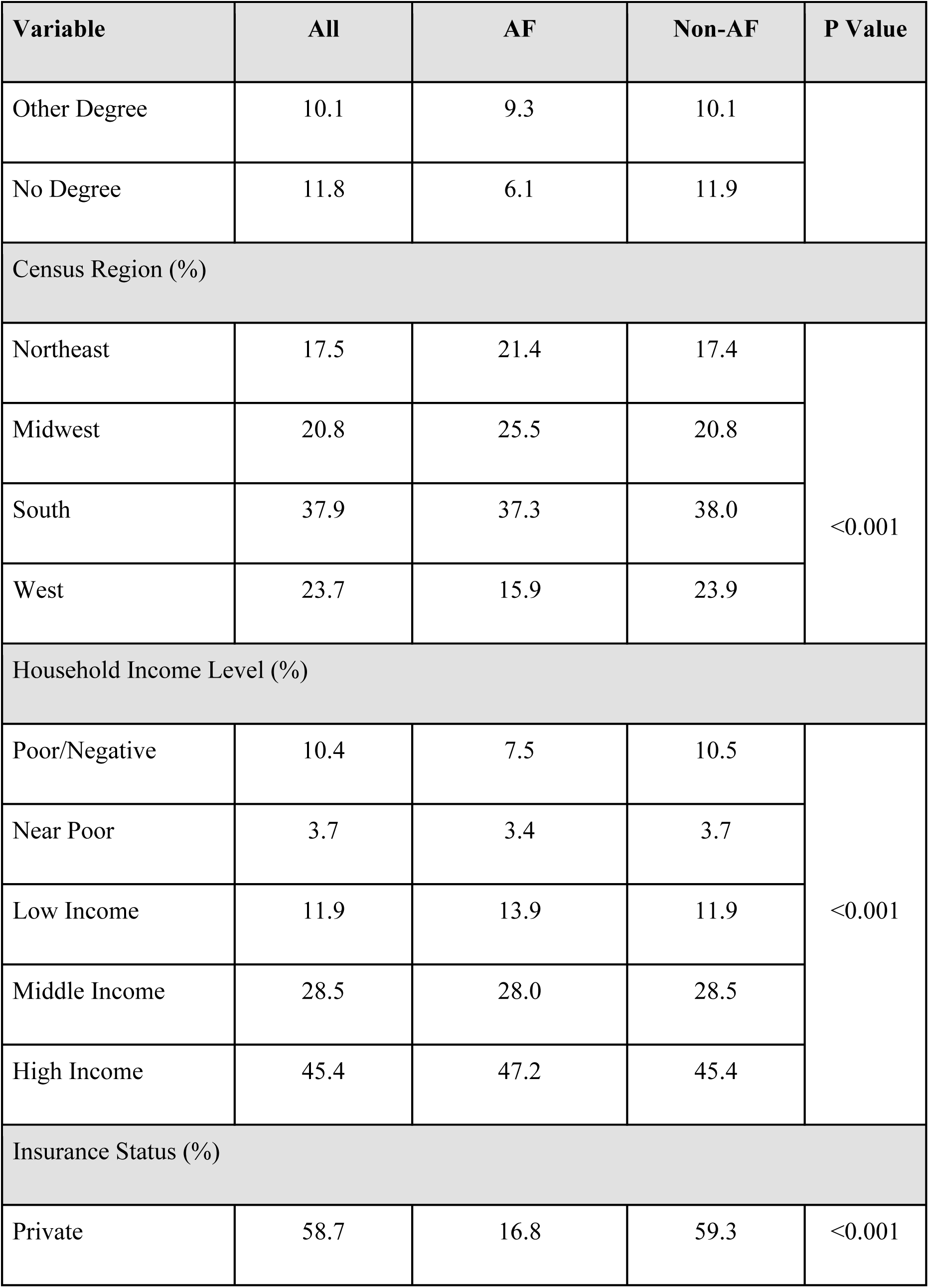
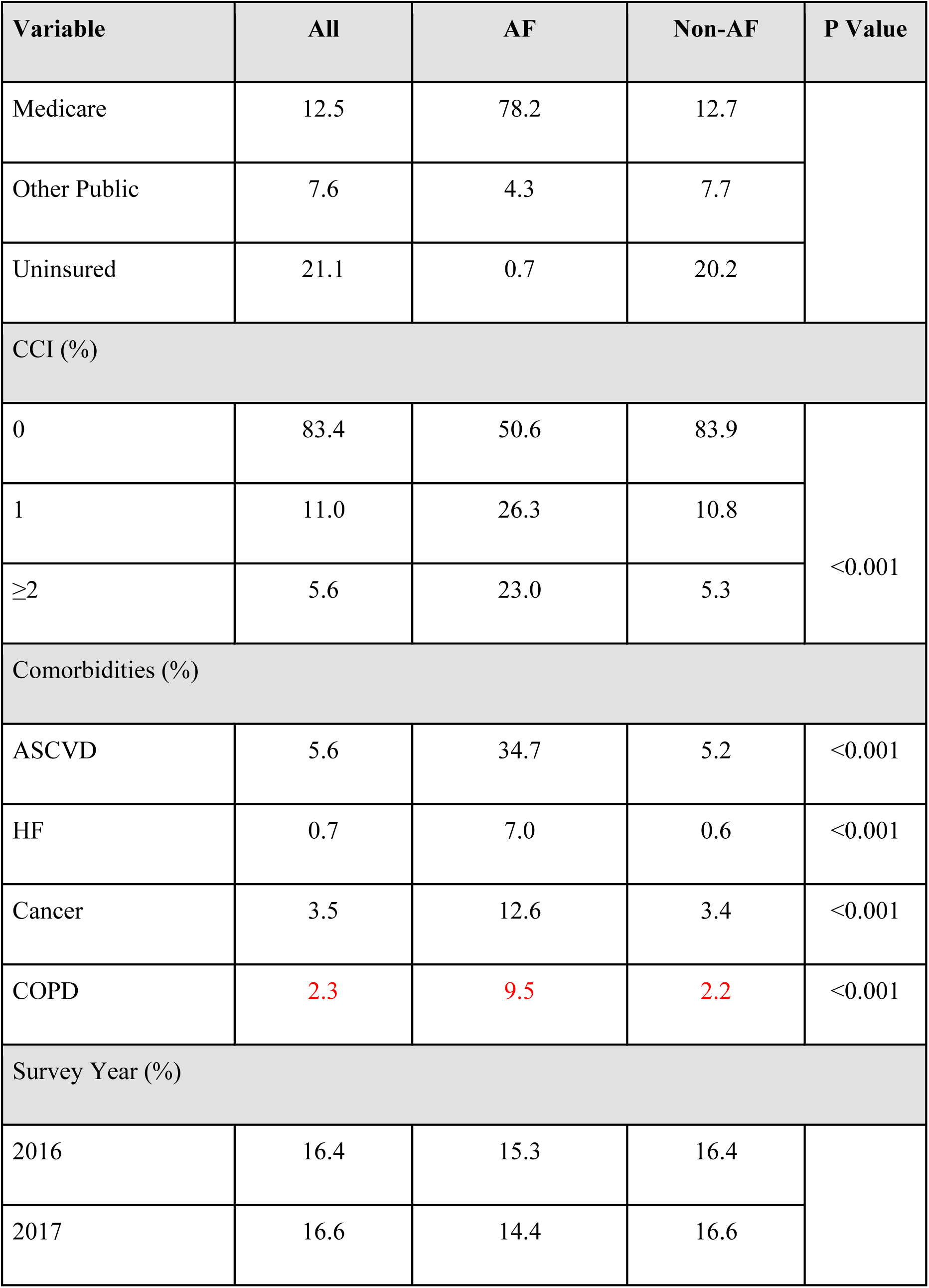
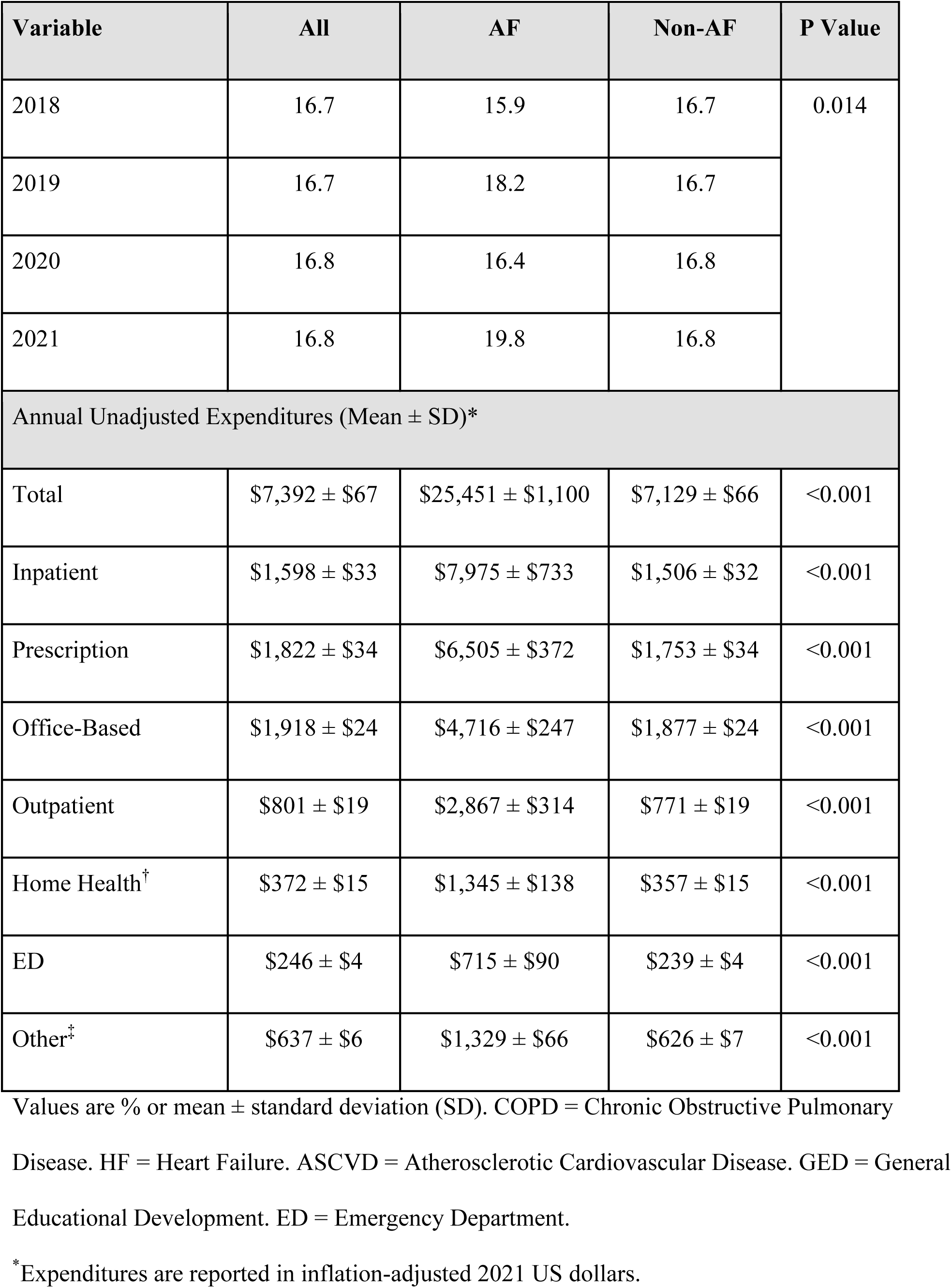
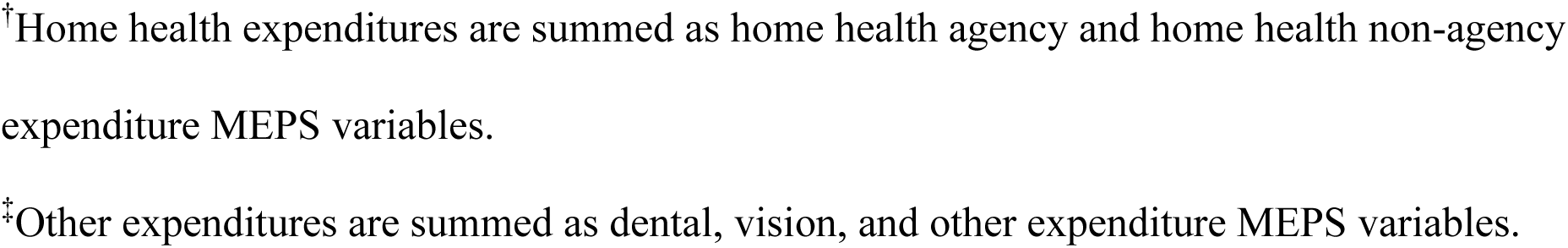
Demographics, Comorbidities, and Annual Expenditures of Individuals with and without AF from 2016-2021.

### Healthcare Expenditures with AF

For individuals with AF, the mean unadjusted annual total healthcare expenditure in terms of 2021 inflation-adjusted dollars was $25,451 ± $1,100 (Table 1) compared with $9,254 ± $82 for individuals without AF (p<0.001). The highest expenditures among service categories in AF were inpatient visits ($7,975 ± $733), followed by prescriptions ($6,505 ± $372) and office-based visits ($4,716 ± $247). Annual expenditures for individuals with AF were higher than those without AF across all service categories.

In comparison, mean unadjusted annual total healthcare expenditure among individuals with other chronic diseases - HF, cancer, ASCVD, and COPD - were $33,753 ± $2,026, $25,212 ± $998, $23,096 ± $530, and $21,725 ± $675 respectively.

Trends in mean unadjusted annual expenditure among individuals with AF over 2016-2021 are shown in Figure 1. Over the study period, there was a 11.1% ($2,635) increase in annual total expenditures, 63.8% ($1,667) increase in outpatient expenditures, 31.6% ($1,193) increase in office-based visits, 97.8% ($493) increase in emergency department expenditures, 9.0% ($133) increase in home health expenditures, and 49.0% ($485) increase in other expenditures. There was a decrease of 14.6% ($1,129) in inpatient expenditures and 3.1% ($208) in medication expenditures. In comparison, among individuals without AF, there was a 23.1% ($1,464) increase in annual total expenditures, driven by a 41.9% ($672) increase in office-based expenditures, 87.0% ($490) increase in outpatient expenditures, 21.1% ($335) increase in medication expenditures, and 48.0% ($228) increase in other expenditures. There was a 13.9% ($214) decrease in inpatient expenditures, 4.9% ($16) decrease in home health, and a 12.4% ($30) decrease in ED expenditures.

**Figure 1:**
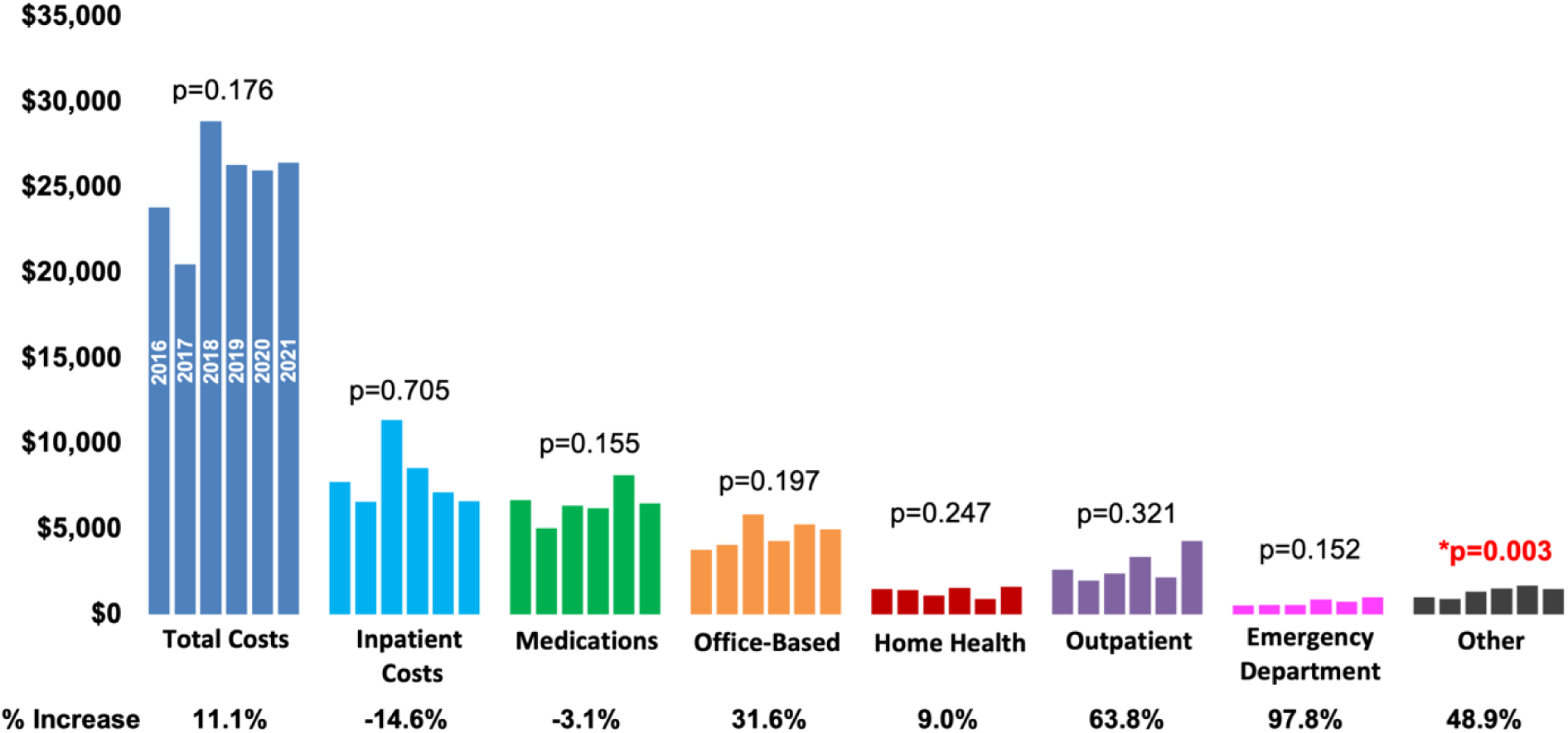
Trends in Healthcare Expenditures over Time for Individuals with AF. This bar graph shows the trends in mean unadjusted annual total healthcare expenditures for individuals with AF broken down by medical service category from 2016 to 2021.

After adjusting for demographics, comorbidities, and survey year, the incremental mean annual healthcare expenditure attributable to AF estimated from the two-part regression model was $6,188 per person (95% CI $5,075-$7,301). With a survey weighted AF population of 3,564,763 as shown in Table 1, this translates to an overall national healthcare expenditure of $22.1 billion per year.

Incremental expenditures attributable to AF within subpopulations of individuals with the 4 other common chronic comorbidities of cancer, ASCVD, COPD, and HF were estimated using a gamma regression model for each subpopulation (Table 2). From largest to smallest, the average marginal effects (95% CI) of having AF in addition to cancer, COPD, or ASCVD were $11,967 ($4,410-$19,525), $8,104 ($2,594 - $13,613), and $6,721 ($3,684 - $9,758) respectively. HF did not significantly impact AF expenditures with an average marginal effect of -$2,756 (-$10,048 -$4,535) (Central Illustration).

**Table 2:**
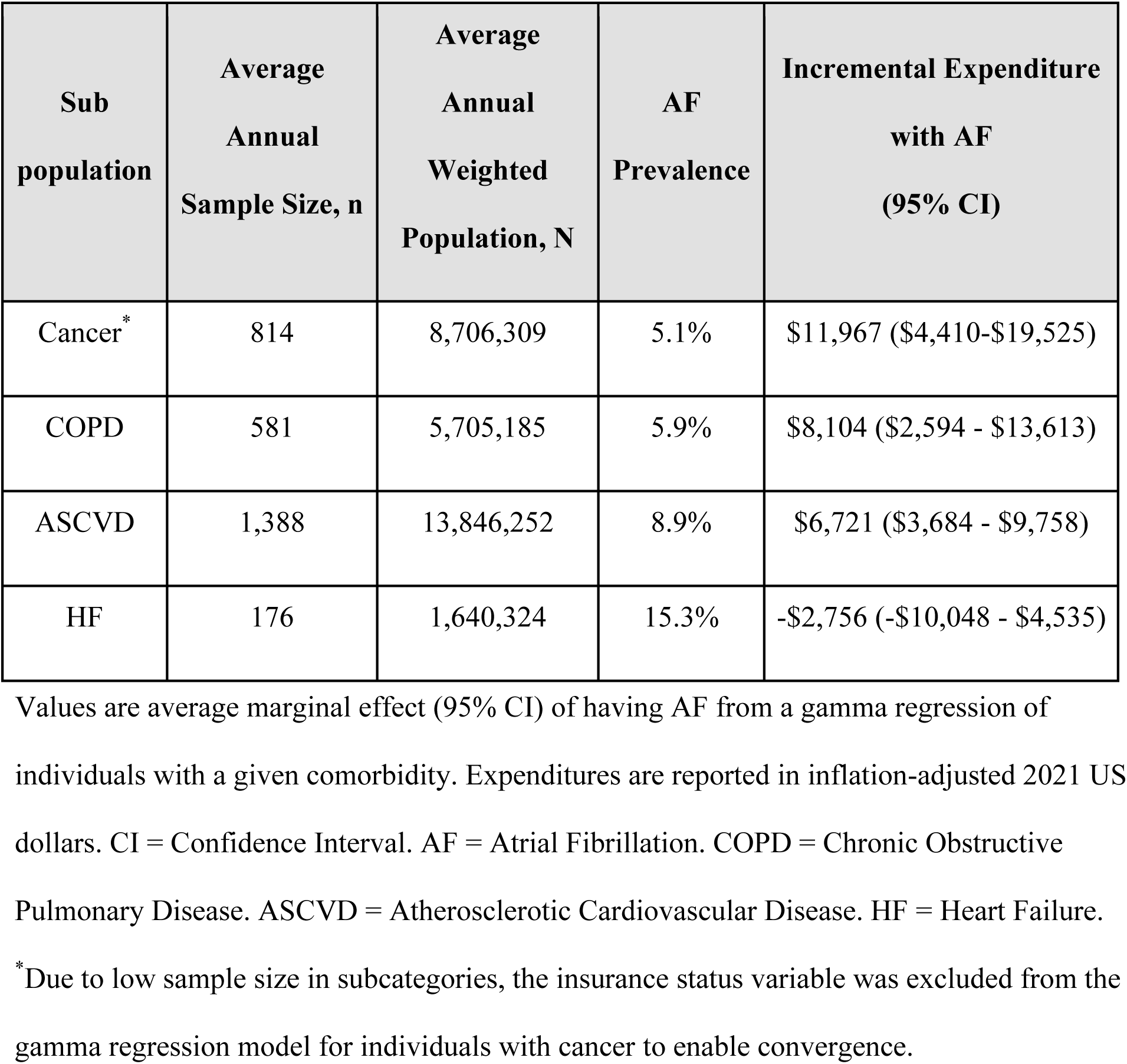
Incremental Expenditures with AF among Individuals with Other Common Chronic Comorbidities from Gamma Regression.

After adjustment for demographics, comorbidities, and survey year in a gamma regression model (Table 3), the variables associated with increased total healthcare expenditure among individuals with AF were modified CCI of ≥2 [OR 1.95 (95% CI 1.53-2.50), p<0.001] and 1 [OR 1.28 (95% CI 1.07-1.54), p=0.009], uninsured status [OR 1.83 (95% 1.13-2.96), p=0.014], cancer [OR 1.76 (95% CI 1.35-2.29), p<0.001], poor income level [OR 1.39 (95% CI 1.02-1.88), p=0.037], ASCVD [OR 1.37 (95% CI 1.17-1.60), p<0.001], COPD [OR 1.31 (95% CI 1.01-1.69), p=0.039], and later survey year [OR 1.07 (95% CI 1.03-1.12), p=0.002].

**Table 3:** Factors Associated with Increased Total Healthcare Expenditures Among Individuals with AF from Gamma Regression.

|  | OR (95% CI) | P Value |
| --- | --- | --- |
| Age Categories |  |  |
| 65-84 years old (Ref) |  |  |
| 18-44 years old | 1.37 (0.35-5.34) | 0.648 |
| 45-64 years old | 0.85 (0.33-2.16) | 0.729 |
| ≥85 years old | 1.10 (0.89-1.34) | 0.381 |
| Sex |  |  |
| Male (Ref) |  |  |
| Female | 0.92 (0.79-1.07) | 0.274 |
| Survey Year | 1.07 (1.03-1.12) | 0.002 |
| Race/Ethnicity |  |  |
| Non-Hispanic White (Ref) |  |  |
| Hispanic | 0.98 (0.73-1.32) | 0.896 |

|  | **OR (95% CI)** | **P Value** |
| --- | --- | --- |
| Non-Hispanic Black | 0.90 (0.63-1.27) | 0.545 |
| Non-Hispanic Asian and Other/Mixed | 1.15 (0.65-2.04) | 0.638 |
| Modified CCI |  |  |
| 0 (Ref) |  |  |
| 1 | 1.28 (1.07-1.54) | 0.009 |
| $\geq 2$ | 1.95 (1.53-2.50) | <0.001 |
| Comorbidities |  |  |
| Cancer | 1.76 (1.35-2.29) | <0.001 |
| ASCVD | 1.37 (1.17-1.60) | <0.001 |
| COPD | 1.31 (1.01-1.69) | 0.039 |
| HF | 1.20 (0.95-1.52) | 0.124 |
| Education |  |  |
| GED/HS Grad (Ref) |  |  |
| No Degree | 0.79 (0.59-1.06) | 0.110 |
| Bachelor's | 1.19 (0.93-1.54) | 0.168 |
| Master/Doctorate | 1.20 (0.95-1.51) | 0.124 |
| Other Degree | 0.83 (0.66-1.03) | 0.096 |
| Marital Status |  |  |
| Married (Ref) |  |  |
| Widowed/ Divorced/ Separated | 1.12 (0.97-1.29) | 0.108 |
| Never Married | 1.04 (0.76-1.43) | 0.798 |
| Household Income Level |  |  |
| High Income (Ref) |  |  |
| Poor | 1.39 (1.02-1.88) | 0.037 |
| Near Poor | 1.11 (0.86-1.45) | 0.398 |
| Low Income | 1.04 (0.84-1.28) | 0.724 |
| Middle Income | 1.14 (0.98-1.33) | 0.099 |
| Insurance Status |  |  |
| Medicare (Ref) |  |  |
| Private | 1.78 (0.66-4.78) | 0.254 |
| Other Public | 1.18 (0.42-3.28) | 0.749 |
| Uninsured | 1.83 (1.13-2.96) | 0.014 |

OR = Odds Ratio. CI = Confidence Interval. CCI = Charlson Comorbidity Index. COPD = Chronic Obstructive Pulmonary Disease. HF = Heart Failure. ASCVD = Atherosclerotic Cardiovascular Disease. AF = Atrial Fibrillation. GED = General Education Development. Grad = Graduate. HS = High School. Ref = Reference.

## DISCUSSION

Individuals with AF had mean unadjusted annual total healthcare expenditures nearly 3 times that of individuals without AF and total expenditures increased by 11% from 2016 to 2021. After adjustment for demographics, comorbidities, and survey year, the incremental annual expenditure was ∼$6,000 per person with AF, translating to over $22 billion in healthcare expenditures annually from AF. Among individuals with AF, a higher comorbidity burden, presence of cancer, uninsured status, and a more recent survey year were associated with higher expenditures.

Our findings build on prior studies focusing on healthcare expenditures in AF. In a systematic review of studies from 1990-2009, Wolowacz et al. found the direct expenditures of AF across the 5 studies from the US to vary from as low as $2,000 per patient-year to as high as $14,200 (23). In another study, using a database of commercially insured patients from 2017-2020, Deshmukh et al. found mean total annual healthcare expenditure for patients with AF to be much higher at $63,031 ($27,896 more than those without AF) (24). Additionally, in a 1999-2002 privately insured database for 2 million enrollees, Wu et al. reported that the excess annual direct expenditure of AF was $12,349 (p<0.001), with individuals with AF having expenditures approximately 5 times higher than individuals without AF ($15,553 versus $3,204, respectively) (25).

In our study, we found mean unadjusted annual healthcare expenditures for individuals with AF to be $25,241 per person per year, and after adjustment for demographics and comorbidities, the presence of AF was associated with an additional $6,188 incremental healthcare expenditure. Additionally, although AF is often considered to be a condition associated with other chronic conditions, AF is also, in itself, a significant driver of expenditures. Healthcare expenditures for individuals with AF were noted to be in a similar range to the total expenditures for individuals with other common comorbidities such as ASCVD, cancer, and COPD. With increasing prevalence of AF and healthcare expenditures comparable to other major chronic conditions, healthcare expenditures for AF deserve greater public attention and policy focus.

Compared to the wide range of expenditures reported by prior studies, our estimates are likely more accurate and representative of current expenditures among individuals with AF in the US as the MEPS is considered a national standard in assessing expenditures of medical conditions in the population, measuring both direct payments for healthcare services and out-of-pocket payments, and includes participants across all insurance statuses.

Inpatient (hospitalization) expenditures were the largest contributor to expenditures among individuals with AF—consistent with prior studies (5,23)—and accounted for nearly a third of the total expenditure. Hospitalizations among individuals with AF are often related to the presence of multiple comorbidities or complications related to AF such as stroke, thromboembolism, and anticoagulant-related issues (24,26). However, use of ablation procedures and cardioversions to maintain sinus rhythm and left atrial appendage for stroke prevention may also be an increasing contributor to hospitalizations. One reason for the increase in total healthcare expenditures from 2016-2021 could be increasing use of catheter ablation strategies for AF patients, which have grown by 10% per year (27). However, studies have suggested that ablation may reduce downstream costs compared to medical therapy. In the Catheter Ablation vs Antiarrhythmic Drug Therapy for Atrial Fibrillation trial (CABANA) trial, catheter ablation of AF was economically attractive compared to drug therapy based on the incremental cost-effectiveness ratio of $57,893 per quality-adjusted life year gained (28). It is possible that over time the increased use of ablations may translate to decreased hospitalization costs. The strength of the ongoing survey design of MEPS can allow periodic reassessment of hospitalization costs. Future studies distinguishing expenditures due to elective vs. emergent hospitalizations can improve our understanding of expenditures related to AF. Ultimately, as hospitalizations contribute significant burden in expenditures with AF (29), they should also be afforded significant policy focus to improve patient care.

Prescription expenditures were the second largest contributor to AF expenditures. A recent innovation in AF management has been the emergence of DOACs and their use has been increasing use of over the past decade (30). Besides DOACs, increasing antiarrhythmic drug usage may also contribute to increasing prescription expenditures (31). Despite high prescription expenditures, our study showed a small decrease in prescription expenditures of 3.1% ($208) from 2016-2021. This may be because most DOACs were already approved prior to our study’s start year of 2016 (32). With some DOACs potentially becoming available in generic form in the future, prescription expenditures may decrease substantially over the next decade. Furthermore, medications such as DOACs reduce severe bleeding events including intracranial bleeding and left atrial appendage occlusion.

Among individuals with AF, the mean unadjusted annual total expenditure increased over time by $2,636, or 11.1%. Over the 2016-2021 study period, there were increases across all medical service categories except for inpatient expenditures (the category with the largest contribution to expenditures), which decreased by 14.6%, and medication expenditures, which decreased by 3.1%. A closer look at the trend in inpatient expenditures shows that there was an increasing trend of hospitalization expenditures prior to the COVID-19 pandemic (22.0% from 2016-2019). However, since then, the inpatient expenditures were observed to decrease by 29.9% from 2019-2021, which match reported declines in AF admission rates and new diagnoses during COVID-19 (33-35). The drivers of this recent decrease are unclear. This may be related to the competing risk of mortality, which was high among the elderly population with COVID-19—especially among those with multiple comorbidities. Alternatively, there may have been systemic declines in hospitalizations or average hospitalization expenditures among individuals with AF. Studies are needed to compare the causes, frequency, and expenditures of hospitalizations among individuals with AF before and after the pandemic.

A novel aspect of our study is the evaluation of the incremental expenditures attributable to AF among individuals with other commonly coexisting chronic diseases. Incremental expenditures attributable to AF were greatest in cancer ($11,967), suggesting that coexistence or development of AF is a significant cost driver among individuals with cancer. This may be related to increased hospitalizations or length of stay and increased prescription expenditures. On the other hand, AF did not change expenditures for individuals with HF, which is surprising given that AF is associated with worse prognosis in HF (36) and can be expected to increase costs. However, a lack of effect on expenditures when these conditions co-exist may potentially be due to an overlap in shared medications, treatments for these two chronic cardiac conditions, or unidentified confounders in baseline characteristics.

In the coming years, the total expenditures for individuals with AF may continue to increase. Newer classes of AF medications and the increasing use of procedural and device therapies may be drivers of expenditures. On the other hand, with more efficacious treatments and monitoring, there may be reductions in other expenditures, such as decreased hospitalizations. The ongoing survey and consistent methodology are strengths of using the MEPS database, and allow for close monitoring of patterns of change in expenditures over time.

Our study has several limitations. First, due to high rates of comorbidities among individuals with AF, there may be residual confounding after multivariable adjustment making it harder to assess independent effects of AF on expenditure. Second, our study overlapped with the COVID-19 pandemic from 2019-2021. Thus, there may be short-term changes in expenditure patterns that warrant reassessment in coming years. Third, because the MEPS database lacks information on procedures, we are unable to report expenditures of AF-associated procedures such as ablation and left atrial appendage closure and how they relate to expenditures as they are likely captured as part of inpatient expenditures. MEPS also lacks participant representation from institutionalized individuals including nursing home residents (37). Also, our study does not account for indirect expenditures incurred by individuals, such as income lost due to reduced productivity and caregiver expenses. Thus, actual expenditures incurred by individuals with AF may be higher than the direct expenditures reported in the study.

An additional limitation is that the self-reported nature of MEPS data can lead to underreporting of expenditures. For personal healthcare spending estimates, MEPS data have been reported to be consistently lower than National Health Expenditure Accounts (NHEA) data from the Centers for Medicare and Medicaid Services, which are known as the official estimates of total healthcare spending in the US. Specifically, MEPS reports $240.3 billion or 17.6% less expenditures than the adjusted NHEA total expenditures (38). These different may be caused by different factors including sampling bias, non-response bias, attrition, and other measurement errors (39).

## CONCLUSION

Individuals with AF have high and increasing healthcare expenditures that amount to over $22 billion in US spending every year. Further, the co-existence of AF among individuals with comorbidities of ASCVD, cancer, and COPD significantly increased expenditures. Given these high expenditures and increasing prevalence, AF warrants increased focus from healthcare policy makers to alleviate financial burden and improve patient outcomes. In parallel, there are continued advancements in AF management, and the resulting improvements in cardiovascular health can lead to reductions in long-term healthcare expenditures. Thus, there is a need to periodically reassess expenditures.

## PERSPECTIVES

### Competency in Systems-Based Practice 1

Individuals with AF incur $25,451 of healthcare expenditures per year and expenditures are increasing over time. The incremental expenditure per year attributable to AF is $6,188 after multivariable adjustment. This translates to an overall national healthcare expenditure of $22.1 billion per year. AF increases expenditures further among individuals with other chronic diseases.

### Competency in Systems-Based Practice 2

Individuals with AF who were surveyed in more recent years, had more comorbidities including cancer, ASCVD, and COPD, and were poor and uninsured had higher annual healthcare expenditures after multivariable adjustment.

### Competency in Patient Care and Procedural Skills

Given high expenditures, AF needs increased focus of policy makers. With increased detection and changes in treatment of AF, expenditures need continual assessments.

### Translational Outlook

This study, which uses an all-payer nationally representative annual cross-sectional survey of medical expenditures of the United States civilian noninstitutionalized population, found that 1) individuals with AF had mean unadjusted annual total healthcare expenditures nearly 3 times that of individuals without AF, with total expenditures increasing by 11.1% from 2016 to 2021; 2) after multivariable adjustment, the incremental annual expenditure was approximately $6,000 per person with AF, translating to over $22 billion in healthcare expenditures annually; and 3) among individuals with AF, a higher comorbidity burden, presence of cancer, ASCVD, or COPD, uninsured status, poor income level, and later survey year were associated with higher expenditures. With an aging US population and new screening and management methods, further studies are needed to reassess trends in healthcare expenditures in AF over time.

## Data Availability

All data and materials are de-identified and publicly available online at https://meps.ahrq.gov/mepsweb/

## Condensed Abstract Abbreviations

AF: Atrial Fibrillation
CCI: Charlson Comorbidity Index
MEPS: Medical Expenditure Panel Survey
NHEA: National Health Expenditure Accounts
US: United States

## Manuscript Abbreviations

AAD: Antiarrhythmic Drug
AF: Atrial Fibrillation
AHRQ: Agency of Healthcare Research and Quality
ASCVD: Atherosclerotic Cardiovascular Disease
CABANA: Catheter Ablation vs Antiarrhythmic Drug Therapy for Atrial Fibrillation
CCI: Charlson Comorbidity Index
COPD: Chronic Obstructive Pulmonary Disease
CPI: Consumer Price Index
DOAC: Direct-Acting Oral Anticoagulants
ED: Emergency Department
FPL: Federal Poverty Line
GED: General Educational Development
HF: Heart Failure
HS: High School
MEPS: Medical Expenditure Panel Survey
NCHS: National Center for Health Statistics
NHEA: National Health Expenditure Accounts
NHIS: National Health Interview Survey
OR: Odds Ratio
SD: Standard Deviation
US: United States
IQR: Interquartile Range

## ACKNOWLEDGEMENTS

We thank the Yale StatLab student consultant team for their statistical advice.

---

**Central Illustration:**
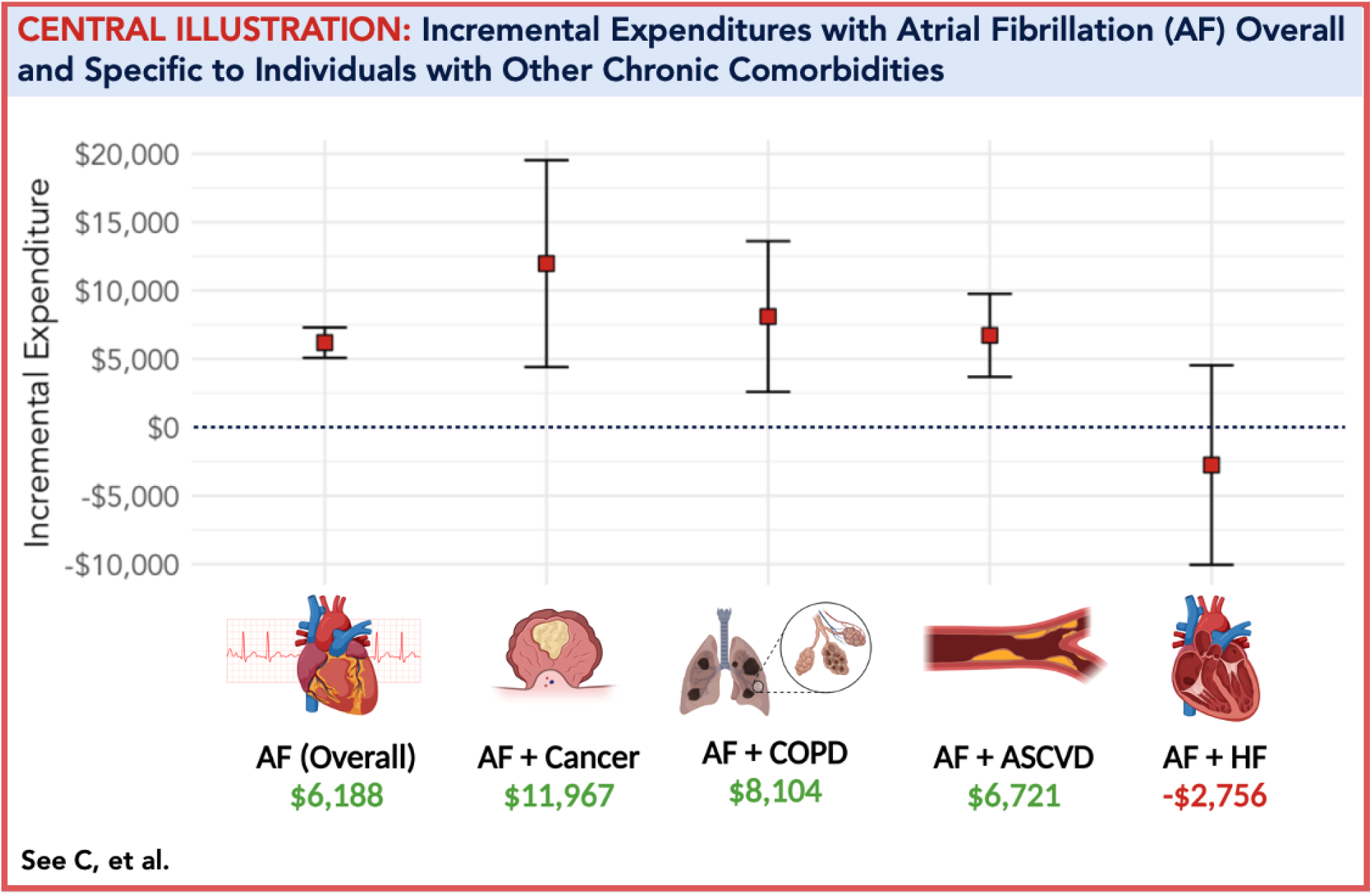
Incremental Expenditures with AF Overall and Specific to Individuals with Other Chronic Comorbidities. This figure shows the incremental expenditures of AF alone and among individuals with cancer, ASCVD, COPD, and HF. Made with BioRender.com (see Supplemental File 1 for publication license). AF = Atrial Fibrillation. ASCVD = Atherosclerotic Cardiovascular Disease. COPD = Chronic Obstructive Pulmonary Disease. HF = Heart Failure.

**Supplemental Table 1:**
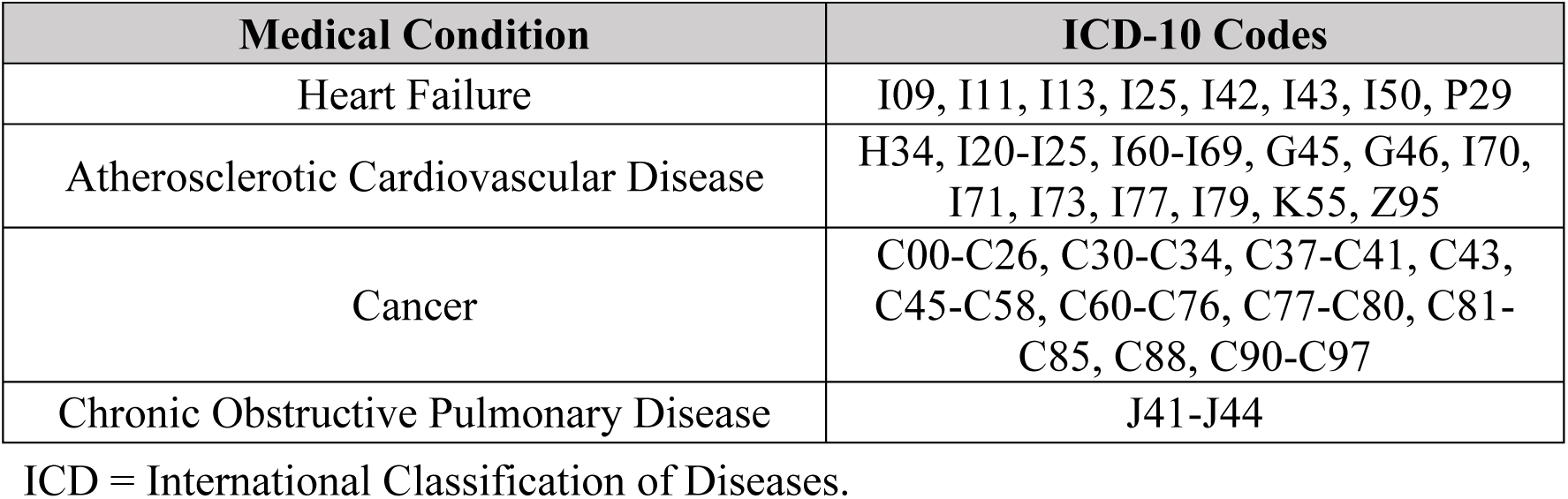
ICD-10 Code Definitions for Other Chronic Comorbidities.

## Notes

### Competing Interest Statement

The authors have declared no competing interest.

### Author Declarations

The source data were openly available prior to the study and continue to be available at: https://meps.ahrq.gov/mepsweb/

